# Maternal over-the-counter analgesics use during pregnancy and adverse perinatal outcomes: Cohort study of 151,141 singleton pregnancies

**DOI:** 10.1101/2020.09.24.20200188

**Authors:** Aikaterini Zafeiri, Edwin A. Raja, Rod T. Mitchell, David C. Hay, Sohinee Bhattacharya, Paul A. Fowler

**Author notes:** **Corresponding Author:** Aikaterini Zafeiri, Institute of Medical Sciences, School of Medicine, Medical Sciences and Nutrition, University of Aberdeen, Aberdeen, United Kingdom.

## Abstract

**OBJECTIVES:** To identify any associations between *in utero* exposure to five over-the-counter (non-prescription) analgesics (paracetamol, ibuprofen, aspirin, diclofenac, naproxen) and adverse neonatal outcomes.

**DESIGN:** Retrospective cohort study using the Aberdeen Maternity and Neonatal Databank.

**PARTICIPANTS:** 151,141 singleton pregnancies between 1985 and 2015.

**MAIN OUTCOME MEASURES:** Premature delivery (<37 weeks), stillbirth, neonatal death, birthweight, standardised birthweight score, neonatal unit admission, APGAR score at 1 and 5 minutes, neural tube and amniotic band defects, gastroschisis and, in males, cryptorchidism, and hypospadias.

**RESULTS:** 83.7% of women taking over-the-counter analgesics reported first trimester use when specifically asked about use at their first antenatal clinic visit. Pregnancies exposed to at least one of the five analgesics were significantly independently associated with increased risks for premature delivery <37 weeks (aOR=1.50, 95%CI 1.43-1.58), stillbirth (aOR=1.33, 95%CI 1.15-1.54), neonatal death (aOR=1.56, 95%CI 1.27-1.93), birthweight <2,500g (aOR=1.28, 95%CI 1.20-1.37), birthweight >4,000g (aOR=1.09, 95%CI 1.05-1.13), admission to neonatal unit (aOR=1.57, 95%CI 1.51-1.64), APGAR score <7 at 1 minute (aOR=1.18, 95%CI 1.13-1.23) and 5 minutes (aOR=1.48, 95%CI 1.35-1.62), neural tube defects (aOR=1.64, 95%CI 1.08-2.47) and hypospadias (aOR=1.27, 95%CI 1.05-1.54 males only). The overall prevalence of over-the-counter analgesics use during pregnancy was 29.1%, however it rapidly increased over the 30-year study period, to include over 60% of women in the last seven years of the study. This makes our findings highly relevant to the wider pregnant population.

**CONCLUSIONS:** Over-the-counter (non-prescription) analgesics consumption during pregnancy was associated with a substantially higher risk for adverse perinatal health outcomes in the offspring. The use of paracetamol in combination with other non-steroidal anti-inflammatory drugs conferred the highest risk. The increased risks of adverse neonatal outcomes associated with non-prescribed, over-the-counter, analgesics use during pregnancy indicate that healthcare guidance for pregnant women regarding analgesic use need urgent updating.

**Funding:** Biotechnology and Biological Sciences Research council (BBSRC) funding under the EASTBIO doctoral training programme (grant number 1942576) to AZ and EU Horizon 2020 project FREIA (Grant Number 825100) to PAF. RTM is supported by MRC Centre for Reproductive Health Grant MR/N022556/1.

## Introduction

Globally 23-85% of women use one or more types of prescribed medications during pregnancy ^1,2^. A similarly high proportion of expectant mothers self-medicate using non-prescription, “over-the-counter” (OTC) medicines ^3,4^ and use during pregnancy is becoming increasingly prevalent, especially in Western countries ^5^. While some analgesics e.g. paracetamol are considered safe to consume throughout pregnancy, use of non-steroidal anti-inflammatory drugs (NSAIDs) is not recommended in pregnancy unless on the advice of a medical specialist and should be avoided beyond gestational week 30 because of the risk of premature closure of the ductus arteriosus. However, current evidence is largely conflicting regarding the safety of gestational analgesic use both for the pregnancy and offspring health ^6^. Several studies have reported increased risks for multiple adverse outcomes including hypospadias, cryptorchidism, amniotic band defects and neural tube defects ^7–11^, whilst others have not found significant associations ^12–17^. Taken overall, this has led to significant concern that postnatal health is adversely affected by maternal analgesic use during pregnancy ^18^.

The use of small cohorts in the current epidemiological studies makes it difficult to draw firm conclusions and definite recommendations^12,17,19,20^. There are other aspects of analgesic use that have to be taken into account. Firstly, due to their abundance, it is not always feasible to determine exact consumption rates and dosage. Secondly, even though the mechanisms of action for most of these compounds is not fully understood, most over-the-counter analgesics can diffuse through the placenta and reach the developing fetus ^21^. Thirdly, maternal pharmacokinetics during pregnancy are altered and there are limited pregnancy safety data for these compounds.

Given the diversity in study population, methodology, sample size and findings in the published studies, we conclude that more extensive data from larger cohorts are essential in order to understand the risks over-the-counter analgesic use during pregnancy pose to neonatal health and function. Here we address many limitations of previous studies by analysing one of the largest cohorts, widest range of health data and, pregnancy use of five over-the-counter analgesics consumed in combination or separately. We report on the prevalence of maternal consumption of five different over-the-counter analgesics during pregnancy and their associations with offspring neonatal outcomes using a large cohort of 151,141 singleton pregnancies spanning three decades of population-based data from a single maternity hospital serving the entire population of Aberdeenshire in the North East of Scotland.

## Materials and Methods

This retrospective cohort study utilised data collected in the Aberdeen Maternity and Neonatal Databank (AMND) in Aberdeen, UK on 151,141 pregnancies over a 30 year period (1985-2015). Details about AMND have been previously published ^22^. Data were collected from medical notes of women retrospectively after delivery. Women were specifically asked about their use of over-the-counter (non-prescription) analgesics at their first antenatal clinic. Data were entered by dedicated coding staff into a computerised database. Data validity was ensured via checking completeness of data entry against NHS (UK National Health Service) returns monthly and constant data cleaning and validation against case notes reported quarterly by the Data Management team to the AMND Steering Committee. A research protocol was submitted and approved by the AMND Steering Committee before data extraction. Approval was received on 6 June 2018. The dataset was fully anonymised, therefore there was no requirement for NHS ethics committee approval.

The main analysis considered consumption during pregnancy of at least one out of five different analgesics: paracetamol (no; yes), ibuprofen (no; yes), naproxen (no; yes), diclofenac (no; yes) or aspirin (no; yes) as the exposure group against no analgesic consumption as the unexposed group. Then, three sub-group analyses against the control group were performed using only paracetamol, only diclofenac, or at least one analgesic from aspirin/naproxen/ibuprofen as exposure groups, excluding pregnancies exposed to multiple analgesics at the same time. As 98.3% of pregnancies using diclofenac were between 2005 and 2015, diclofenac sub-group analysis only considered pregnancies during that time frame in order to rule out any temporal effect.

The offspring outcomes compared between control and exposed groups were: gestation at delivery (preterm <37 gestation weeks, term ≥37 gestation weeks), pregnancy outcome (livebirth, stillbirth, neonatal death), baby weight (low birth weight (LBW) ≤2,499 g, high birth weight (HBW) ≥4,000 g, normal birth weight (NBW) 2,500g-3,999 g), standardised birthweight score was considered as a continuous variable as previously described by Campbell and colleagues^23^, baby admission to neonatal unit (no; yes), APGAR score at one and five minutes (<7, ≥7), cryptorchidism (no; yes) (ICD-10 code Q53), neural tube defects (no; yes) (ICD-10 code Q00-07), amniotic band defects (no; yes) (ICD-10 codes Q70-74), hypospadias (no; yes) (ICD-10 code Q54), gastroschisis (no; yes) (ICD-10 code Q79.3). A composite outcome (presence of at least one congenital anomaly (no; yes)) was created using the variables neural tube defects, amniotic band defects, and gastroschisis and, in males, cryptorchidism and hypospadias.

The baseline characteristics compared between exposed and unexposed pregnancies were (reference category first): year of delivery (1985-1994, 1995-2004, 2005-2015), maternal age at delivery (20-25, <20, 26-35, >35 years), previous pregnancy (no; yes), maternal body mass index (BMI) (normal weight 18.5-24.9 kg/m^2^, underweight <18.5 kg/m^2^, overweight 25-29.9 kg/m^2^, obese <30 kg/m^2^), maternal first antenatal visit (1st, 2nd, 3rd trimester), maternal smoking status (non-smoker, smoker, ex-smoker), Scottish Index of Multiple Deprivation (SIMD) decile (1-6, 7-10, decreasing deprivation with increasing score), maternal hypertensive disorders (no disorder, gestational hypertension, preeclampsia, eclampsia), maternal antepartum haemorrhage (no haemorrhage, abruption, placental previa), type of labour (spontaneous, elective caesarean section, induced), type of delivery (spontaneous vaginal delivery, instrumental, caesarean section), analgesia during labour (no; yes), baby presentation at delivery (occiput anterior, occiput posterior), baby sex (female; male).

## Statistical Analysis

Baseline characteristics were compared between exposed and unexposed pregnancies to any analgesic using χ^2^ test for categorical variables and t-test for normally distributed continuous variables as appropriate. Relationships between exposures and outcomes were examined by binary logistic regression for binary outcome variables, multinomial logistic regression for nominal categorical outcome variables, and multiple linear regression for continuous variables. The strength of association was reported as odds ratios (ORs) with 95% confidence intervals (CI). The socio-demographic characteristics that were likely to confound our exposure-to-outcome path were identified using a directed acyclic graph (DAG) (Figure S1)^24^. Factors that were associated with consumption of over-the-counter analgesics during pregnancy at 10% level of significance and deemed clinically relevant, were included in the model as confounders. All outcomes were adjusted for year of delivery, maternal age at delivery, SIMD and maternal first antenatal visit. In addition to these confounders, individual outcomes were adjusted for relevant cofactors. Gestation at delivery and pregnancy outcome were both additionally adjusted for maternal hypertensive disorders and antepartum haemorrhage. Weight of the baby, neonatal unit admission, cryptorchidism, neural tube defects, amniotic band defects, hypospadias and gastroschisis variables were also adjusted for gestation at delivery. APGAR score at one and five minutes were adjusted for type of delivery. A p-value of less than 0.05 was considered statistically significant. All statistical analyses were carried out using IBM SPSS Statistics version 25.0 (Released 2017. IBM SPSS Statistics for Windows, Armonk, NY: IBM Corp.). R version 3.6.2 was used to generate Figure 2.

## Results

83.7% of women taking over-the-counter analgesics reported first trimester use when specifically asked about use at their first antenatal clinic. Overall, from the total 151,141 pregnancies across 30 years in 107,143 (70.9%) pregnancies, no over-the-counter analgesic consumption was reported. At least one over-the-counter analgesic was consumed in 43,998 (29.1%) pregnancies, whereas paracetamol use alone was reported in 24,099 (18.4%) pregnancies. Diclofenac use was observed in 20.0% of pregnancies in the 10-year period when diclofenac was available over-the-counter (without prescription). Finally, at least one out of three analgesics (naproxen, ibuprofen, aspirin) was consumed in 762 (0.7%) pregnancies (Figure 1).

**Figure 1.**
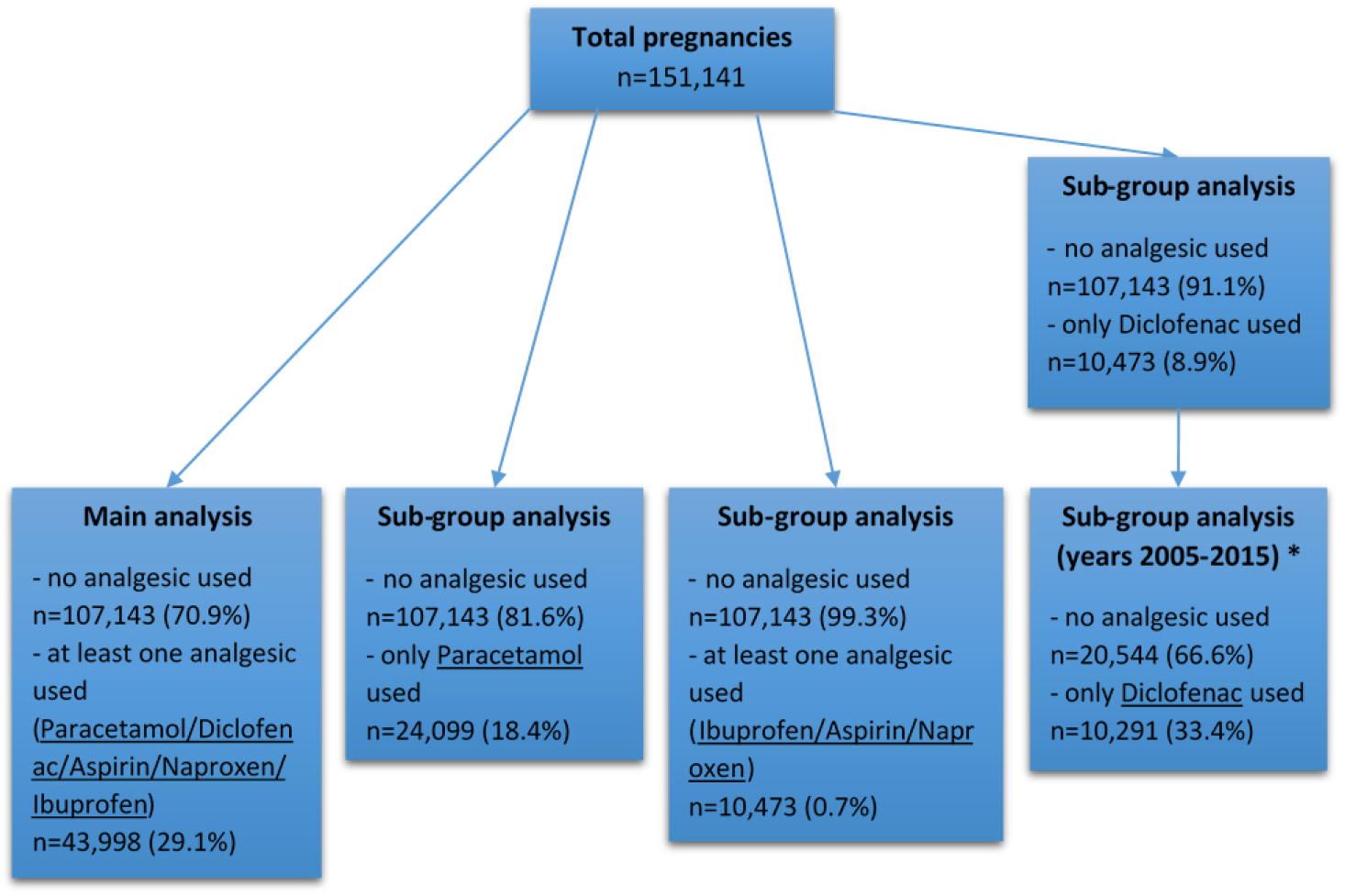
Flowchart of cohort selection and sub-group analyses. n=number of pregnancies in each analysis.* 98.3% of pregnancies using only diclofenac occurred during 2005-2015, therefore analysis was performed only on data from that decade to rule out any temporal effect.

Prevalence of use for all five analgesics increased dramatically over the 30-year study period (1985-2015) (Figure 2). Percentage of pregnancies with consumption of at least one analgesic increased from 1.8% in 1985 to 70.6% in 2015. Paracetamol was consumed in 1.3% of pregnancies in 1985 and it continuously increased reaching 42.2% in 2015. Naproxen, ibuprofen or aspirin consumption during pregnancy was less prevalent (Figure 2A), however it also increased during the 30-year study period, starting at 0.5% in 1985 and reaching 1.9% in 2015 (Figure 2B). Diclofenac was consumed in very few pregnancies between 1985 (<0.01%) and 2005 (0.2%). Percentage of consumption, however, dramatically increased during the next decade following deregulation of diclofenac, reaching 25.0% in just one year (2006) and 45.6% of all pregnancies in 2015.

**Figure 2.**
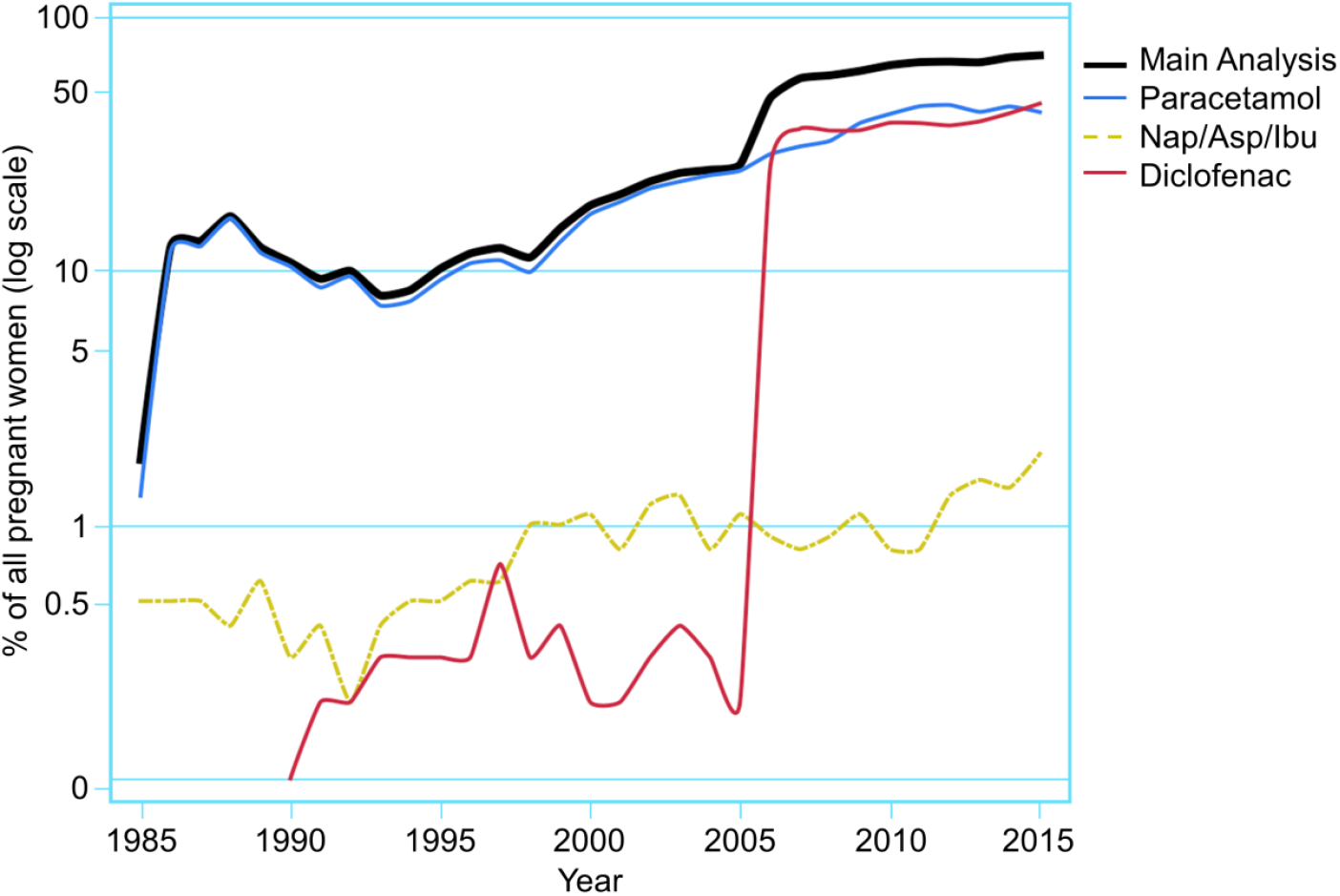
Prevalence of use during pregnancy for each analgesic sub-group over our 30-year study period. In 2005 there was a change in legislation making diclofenac available without prescription.

Table 1 compares the baseline characteristics between the unexposed group of pregnancies where no analgesic was consumed and each of the exposure groups. In most, but not all, comparisons across all four analyses, there was a statistically significant difference (p<0.001) for most variables. In the paracetamol sub-group analysis, baby presentation at delivery (p=0.525) and sex of the baby (p=0.861) were not significantly different between the groups. In the analysis considering consumption of at least one analgesic from aspirin/naproxen/ibuprofen, again the variables for baby presentation at delivery (p=0.093) and sex of the baby (p=0.732), together with maternal smoking status (p=0.132) and maternal antepartum haemorrhage (p=0.434) were not statistically different compared to the unexposed group. All variables were statistically different between unexposed and exposed groups for the main analysis and diclofenac sub-group analysis.

**Table 1.**
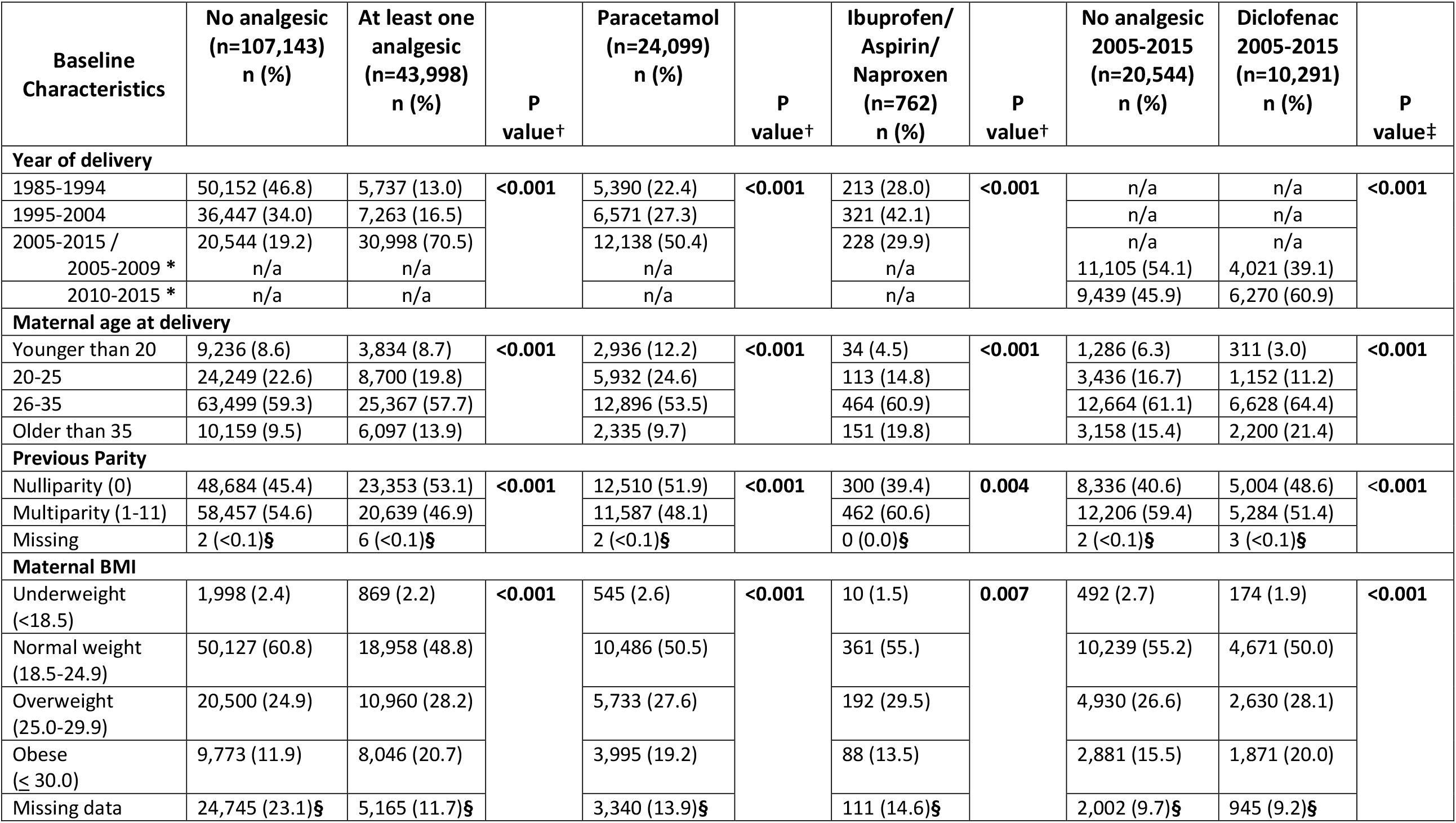
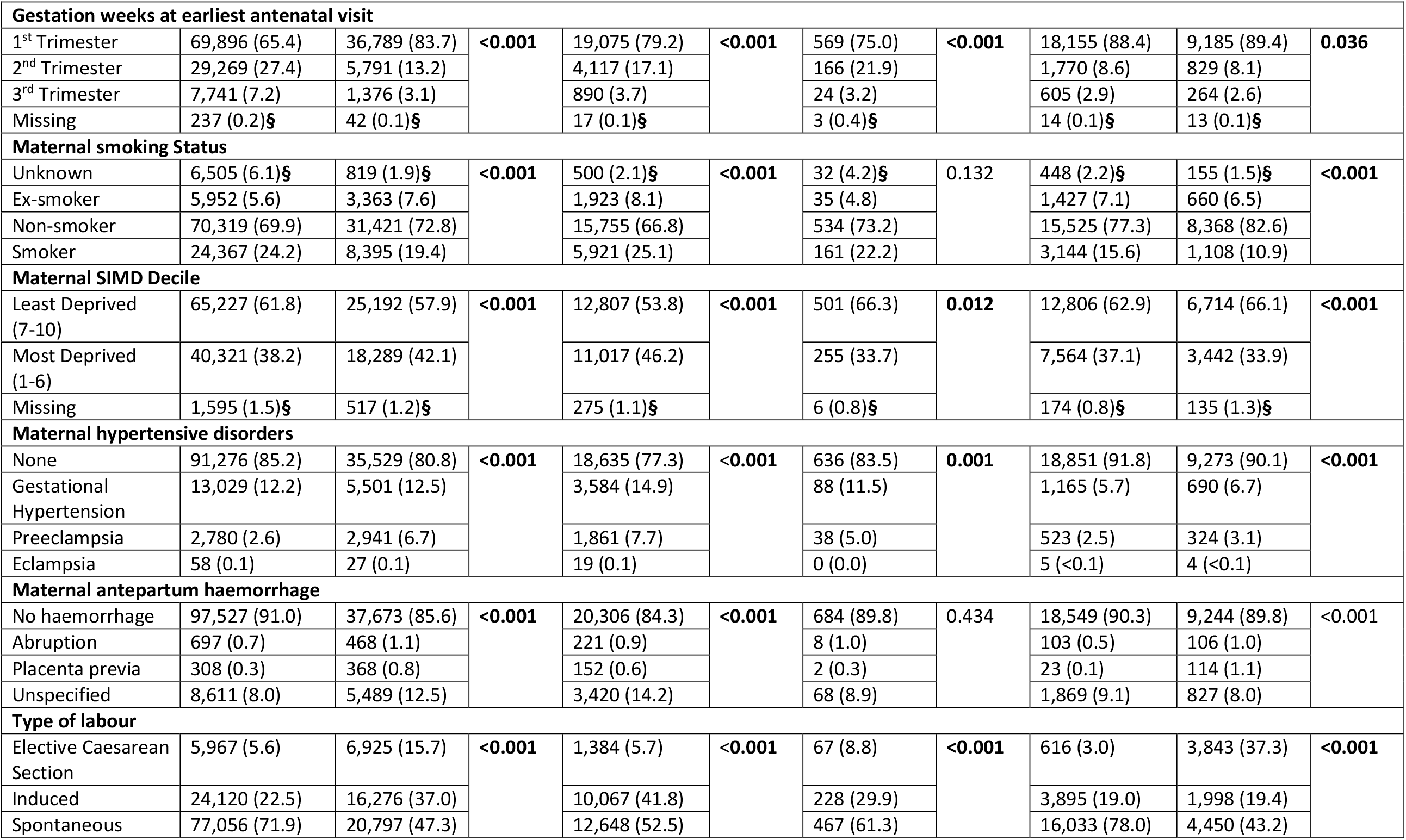
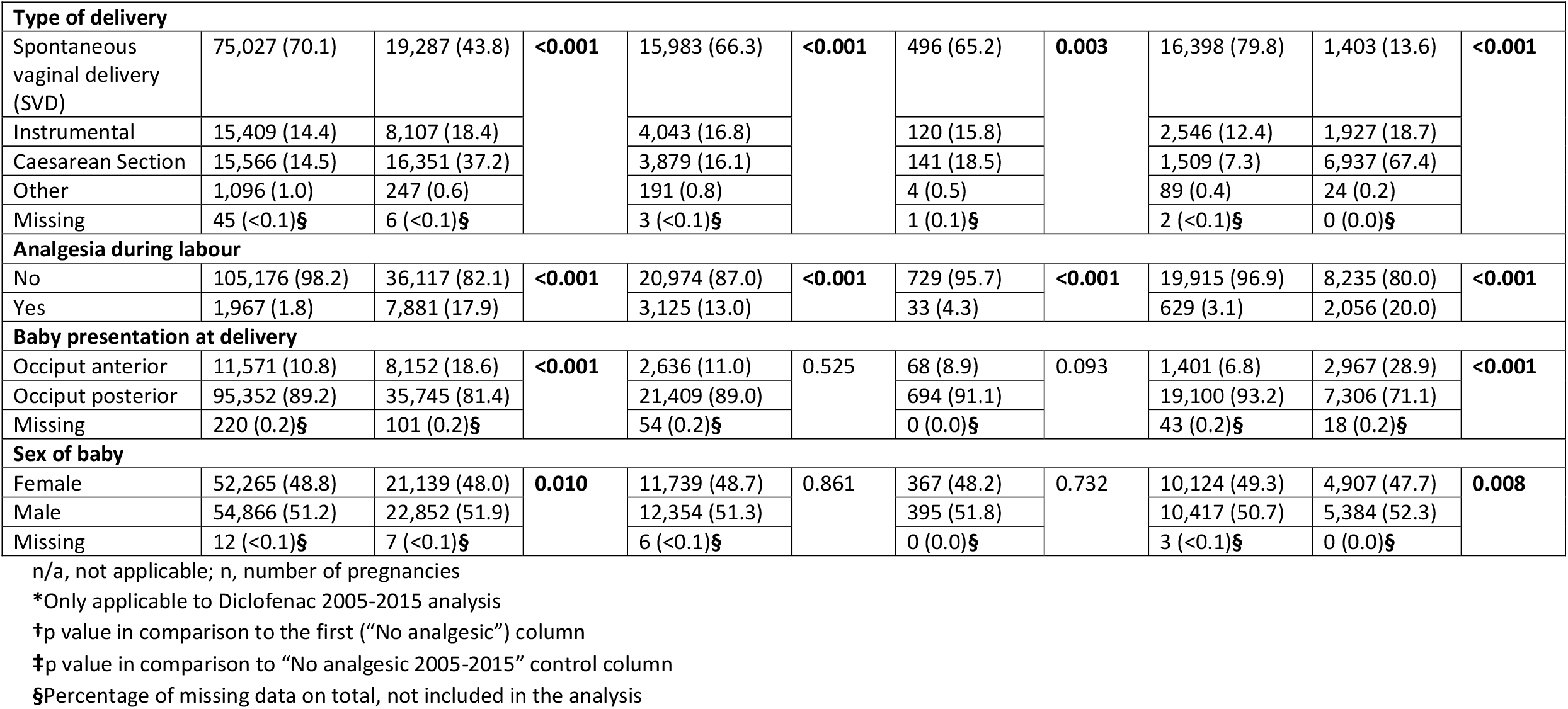
Comparison of baseline characteristics between exposed (use of analgesics) and unexposed (no analgesic use) groups of pregnancies (P values <0.05 shown in bold).

Table 2 summarises the comparison of neonatal outcomes between the unexposed group (no analgesic at all) and the exposed groups of at least one analgesic, only paracetamol and at least one out of aspirin/naproxen/ibuprofen. Comparison of outcomes for the diclofenac sub-group analysis is shown in Table 3.

**Table 2.**
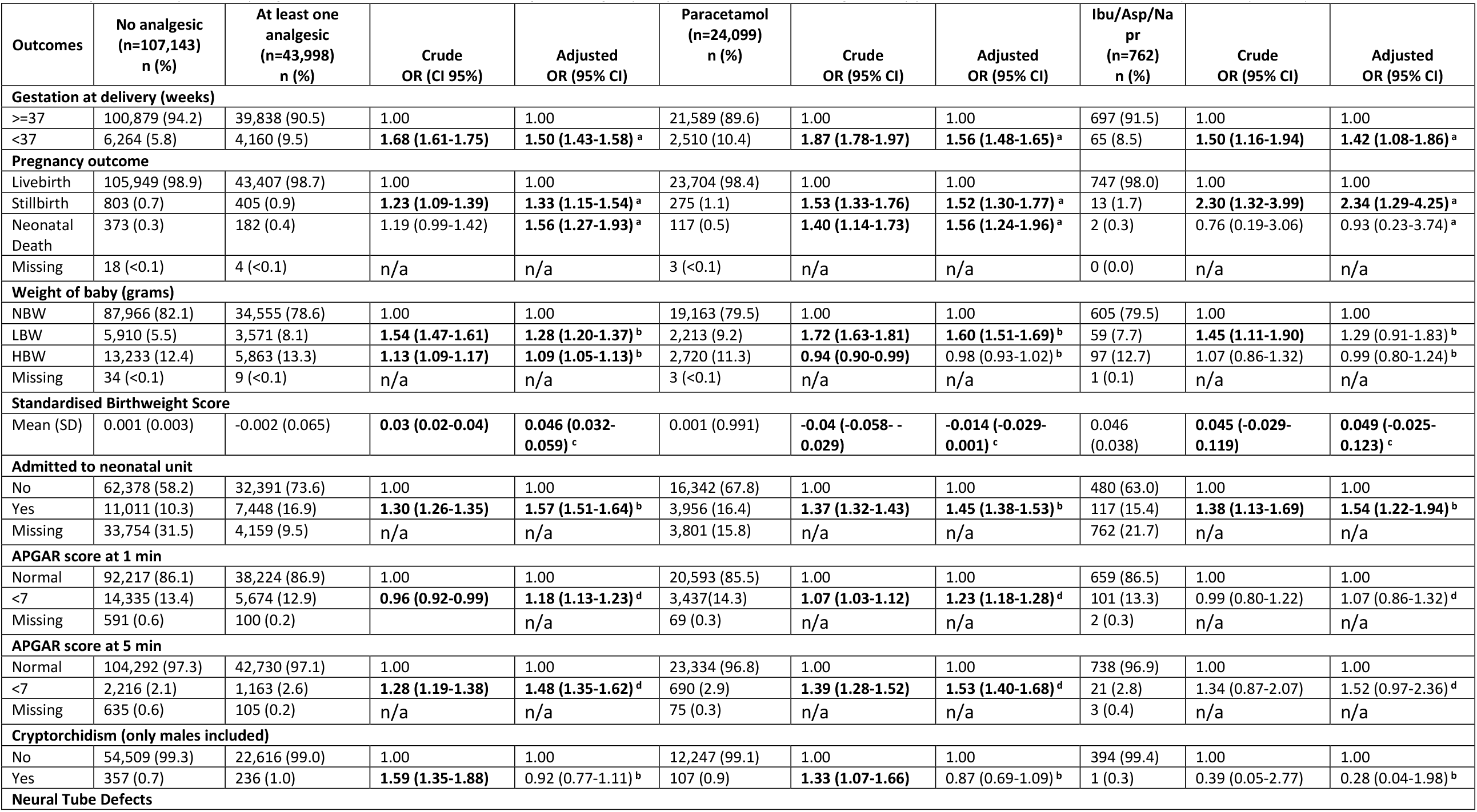
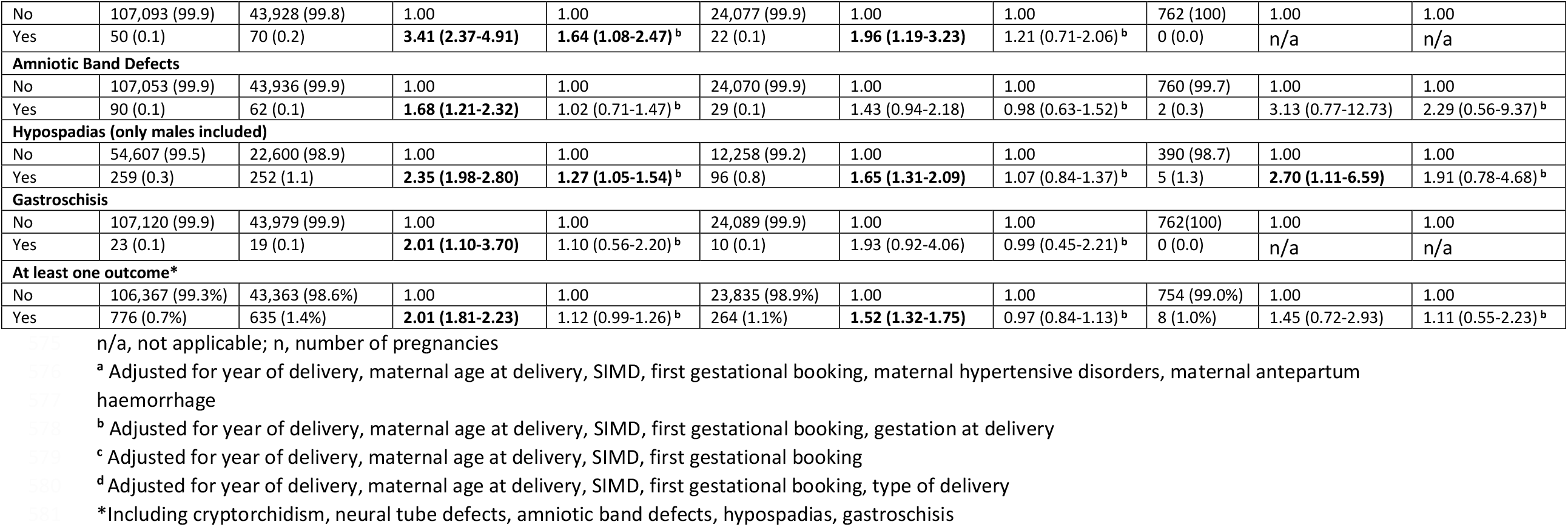
Regression analysis of offspring outcomes between control (no analgesic) and groups exposed to at least one analgesic, only paracetamol, and at least one from ibuprofen, aspirin, naproxen

**Table 3.**
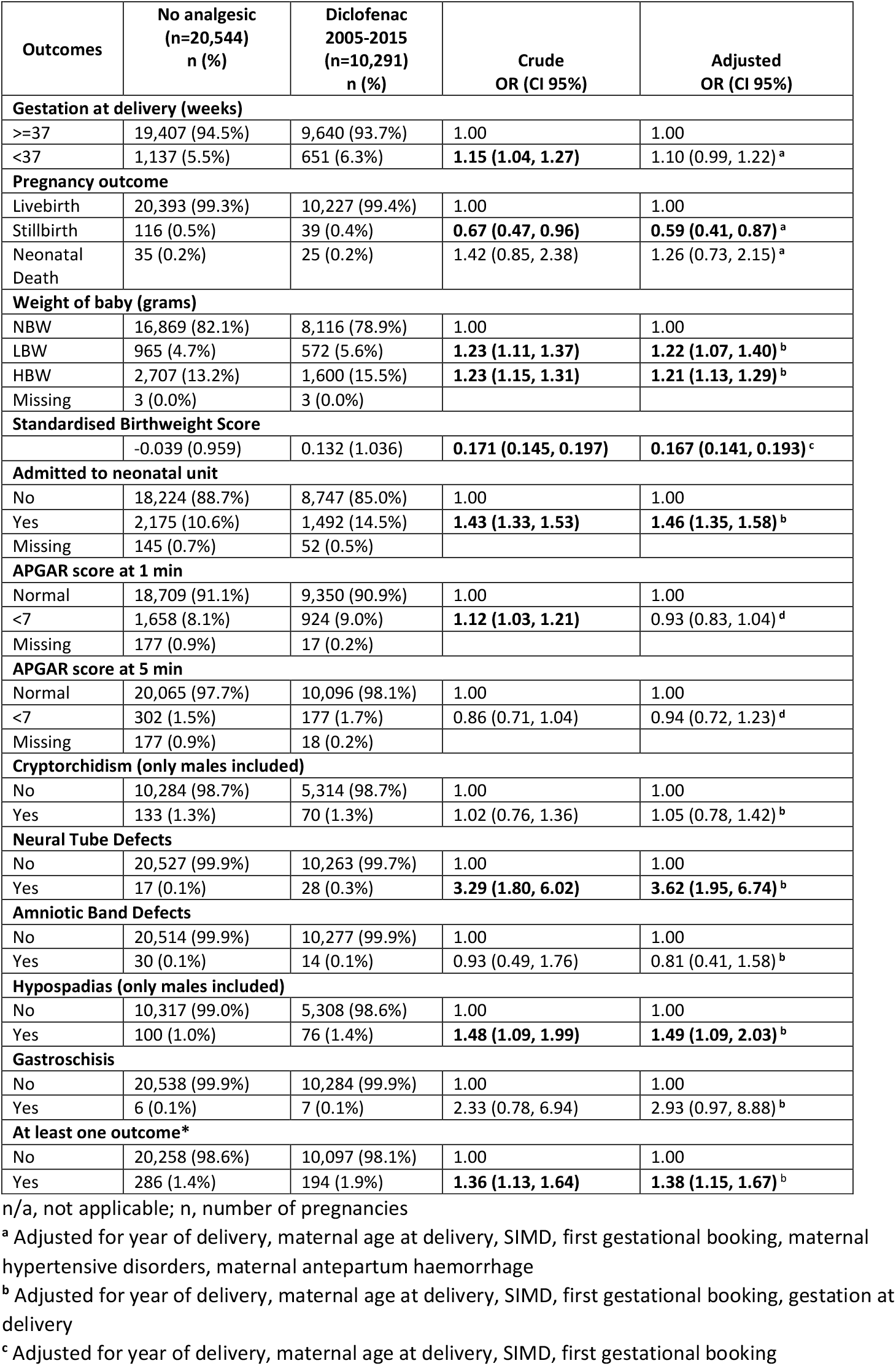
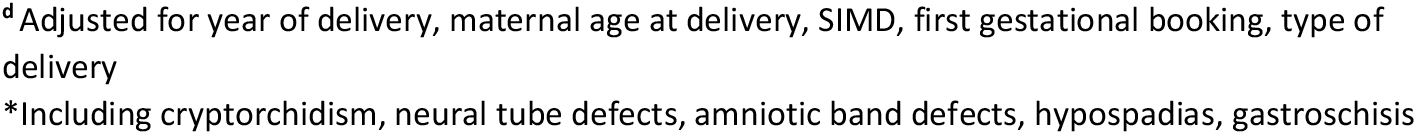
Sub-group regression analysis between control pregnancies and exposed to diclofenac.

| Outcomes | No analgesic<br>(n=20,544)<br>n (%) | Diclofenac<br>2005-2015<br>(n=10,291)<br>n (%) | Crude<br>OR (CI 95%) | Adjusted<br>OR (CI 95%) |
| --- | --- | --- | --- | --- |
| **Gestation at delivery (weeks)** |  |  |  |  |
| ≥37 | 19,407 (94.5%) | 9,640 (93.7%) | 1.00 | 1.00 |
| <37 | 1,137 (5.5%) | 651 (6.3%) | **1.15 (1.04, 1.27)** | 1.10 (0.99, 1.22)<sup>a</sup> |
| **Pregnancy outcome** |  |  |  |  |
| Livebirth | 20,393 (99.3%) | 10,227 (99.4%) | 1.00 | 1.00 |
| Stillbirth | 116 (0.5%) | 39 (0.4%) | **0.67 (0.47, 0.96)** | **0.59 (0.41, 0.87)<sup>a</sup>** |
| Neonatal Death | 35 (0.2%) | 25 (0.2%) | 1.42 (0.85, 2.38) | 1.26 (0.73, 2.15)<sup>a</sup> |
| **Weight of baby (grams)** |  |  |  |  |
| NBW | 16,869 (82.1%) | 8,116 (78.9%) | 1.00 | 1.00 |
| LBW | 965 (4.7%) | 572 (5.6%) | **1.23 (1.11, 1.37)** | **1.22 (1.07, 1.40)<sup>b</sup>** |
| HBW | 2,707 (13.2%) | 1,600 (15.5%) | **1.23 (1.15, 1.31)** | **1.21 (1.13, 1.29)<sup>b</sup>** |
| Missing | 3 (0.0%) | 3 (0.0%) |  |  |
| **Standardised Birthweight Score** |  |  |  |  |
|  | -0.039 (0.959) | 0.132 (1.036) | **0.171 (0.145, 0.197)** | **0.167 (0.141, 0.193)<sup>c</sup>** |
| **Admitted to neonatal unit** |  |  |  |  |
| No | 18,224 (88.7%) | 8,747 (85.0%) | 1.00 | 1.00 |
| Yes | 2,175 (10.6%) | 1,492 (14.5%) | **1.43 (1.33, 1.53)** | **1.46 (1.35, 1.58)<sup>b</sup>** |
| Missing | 145 (0.7%) | 52 (0.5%) |  |  |
| **APGAR score at 1 min** |  |  |  |  |
| Normal | 18,709 (91.1%) | 9,350 (90.9%) | 1.00 | 1.00 |
| <7 | 1,658 (8.1%) | 924 (9.0%) | **1.12 (1.03, 1.21)** | 0.93 (0.83, 1.04)<sup>d</sup> |
| Missing | 177 (0.9%) | 17 (0.2%) |  |  |
| **APGAR score at 5 min** |  |  |  |  |
| Normal | 20,065 (97.7%) | 10,096 (98.1%) | 1.00 | 1.00 |
| <7 | 302 (1.5%) | 177 (1.7%) | 0.86 (0.71, 1.04) | 0.94 (0.72, 1.23)<sup>d</sup> |
| Missing | 177 (0.9%) | 18 (0.2%) |  |  |
| **Cryptorchidism (only males included)** |  |  |  |  |
| No | 10,284 (98.7%) | 5,314 (98.7%) | 1.00 | 1.00 |
| Yes | 133 (1.3%) | 70 (1.3%) | 1.02 (0.76, 1.36) | 1.05 (0.78, 1.42)<sup>b</sup> |
| **Neural Tube Defects** |  |  |  |  |
| No | 20,527 (99.9%) | 10,263 (99.7%) | 1.00 | 1.00 |
| Yes | 17 (0.1%) | 28 (0.3%) | **3.29 (1.80, 6.02)** | **3.62 (1.95, 6.74)<sup>b</sup>** |
| **Amniotic Band Defects** |  |  |  |  |
| No | 20,514 (99.9%) | 10,277 (99.9%) | 1.00 | 1.00 |
| Yes | 30 (0.1%) | 14 (0.1%) | 0.93 (0.49, 1.76) | 0.81 (0.41, 1.58)<sup>b</sup> |
| **Hypospadias (only males included)** |  |  |  |  |
| No | 10,317 (99.0%) | 5,308 (98.6%) | 1.00 | 1.00 |
| Yes | 100 (1.0%) | 76 (1.4%) | **1.48 (1.09, 1.99)** | **1.49 (1.09, 2.03)<sup>b</sup>** |
| **Gastroschisis** |  |  |  |  |
| No | 20,538 (99.9%) | 10,284 (99.9%) | 1.00 | 1.00 |
| Yes | 6 (0.1%) | 7 (0.1%) | 2.33 (0.78, 6.94) | 2.93 (0.97, 8.88)<sup>b</sup> |
| **At least one outcome\*** |  |  |  |  |
| No | 20,258 (98.6%) | 10,097 (98.1%) | 1.00 | 1.00 |
| Yes | 286 (1.4%) | 194 (1.9%) | **1.36 (1.13, 1.64)** | **1.38 (1.15, 1.67)<sup>b</sup>** |

n/a, not applicable; n, number of pregnancies
<sup>a</sup> Adjusted for year of delivery, maternal age at delivery, SIMD, first gestational booking, maternal hypertensive disorders, maternal antepartum haemorrhage585 <sup>b</sup> Adjusted for year of delivery, maternal age at delivery, SIMD, first gestational booking, gestation at delivery587 <sup>c</sup> Adjusted for year of delivery, maternal age at delivery, SIMD, first gestational booking
<sup>d</sup>Adjusted for year of delivery, maternal age at delivery, SIMD, first gestational booking, type of delivery \*Including cryptorchidism, neural tube defects, amniotic band defects, hypospadias, gastroschisis

### All analgesics and neonatal outcomes

As shown in Table 2, compared to unexposed pregnancies in which women did not use any analgesic, pregnancies with consumption of at least one analgesic (paracetamol, diclofenac, aspirin, naproxen, ibuprofen) were independently associated with significantly higher odds for premature delivery (aOR=1.50, 95%CI 1.43-1.58), stillbirth (aOR=1.33, 95%CI1.15-1.54), LBW (aOR=1.28, 95%CI 1.20-1.37), HBW (aOR=1.09, 95%CI 1.05-1.13), baby admission to neonatal unit (aOR=1.57, 95%CI 1.51-1.64), APGAR score <7 at five minutes (aOR=1.48, 95%CI 1.35-1.62), neural tube defects (aOR=1.64, 95%CI 1.08-2.47) and hypospadias (aOR=1.27, 95%CI 1.05-1.54) in adjusted analyses. Significantly decreased odds for APGAR score <7 at one minute were found in the crude analysis (cOR=0.96, 95%CI 0.92-0.99), however when adjusted for year of delivery, maternal age at delivery, SIMD, first gestational booking and type of delivery, the significance changed direction showing significantly increased odds (aOR=1.18, 95%CI 1.13-1.23). A significantly lower standardised birthweight score (aOR=0.046. 95%CI 0.032-0.059) was found for the exposure group compared to no analgesic at all. Cryptorchidism (aOR=0.92, 95%CI 0.77-1.11), amniotic band defects (aOR=1.02, 95%CI 0.71-1.47), gastroschisis (aOR=1.10, 95%CI 0.56-2.20) and the composite outcome variable (aOR=1.12, 95%CI 0.99-1.26), were all associated with increased odds in the exposure group compared to not exposed, however the association was not significant in the adjusted model. There was no significant association between neonatal death and exposure to at least one analgesic in the crude analysis (cOR=1.19, 95%CI 0.99-1.42), however there were significantly higher odds of neonatal death in the adjusted analysis (aOR=1.56, 95%CI 1.27-1.93) in the exposed group compared to control.

### Paracetamol and neonatal outcomes

In the sub-group analysis considering only paracetamol consumption during pregnancy as our exposure group, most of the associations reported in the main analysis remained significant with the same direction of significance (Table 2). The differences were: maternal paracetamol consumption during pregnancy was associated with significantly decreased odds for offspring HBW (cOR=0.94, 95%CI 0.90-0.99) in the crude analysis however significance was lost in the adjusted model (aOR=0.98, 95%CI 0.93-1.02), and there were no significant associations in the adjusted models for neural tube defects (aOR=1.21, 95%CI 0.71-2.06) and hypospadias (aOR=1.07, 95%CI 0.84-1.37).

### Aspirin/naproxen/ibuprofen and neonatal outcomes

Consumption of at least one analgesic from aspirin, naproxen or ibuprofen during pregnancy was compared against the same control group of pregnancies where no analgesic was used (Table 2). Again, when comparing associations between groups in this sub-group analysis and main analysis, fewer outcome variants showed similar significance pattern. The only shared significant associations were for increased odds for premature delivery (aOR=1.42, 95%CI 1.08-1.86), stillbirth (aOR=2.34, 95%CI 1.29-4.25) and baby admission to neonatal unit (aOR=1.54, 95%CI 1.22-1.94) in the adjusted regression analyses.

### Diclofenac and neonatal outcomes

In the sub-group analysis of pregnancies coinciding with non-prescription, over-the-counter, availability of diclofenac (years 2005-2015) were considered, and outcomes compared between the diclofenac group and no analgesic consumption group (Table 3). Compared to the main analysis, diclofenac consumption during pregnancy was not significantly associated with premature delivery (aOR=1.10, 95%CI 0.99-1.22), neonatal death (aOR=1.26, 95%CI 0.73-2.15) and APGAR score <7 in one minute (aOR=0.93, 95%CI 0.83-1.04) in the adjusted models. Associations with APGAR score <7 in five minutes (aOR=0.94, 95%CI 0.72-1.23), cryptorchidism (aOR=1.05, 95%CI 0.78-1.42), amniotic band defects (aOR=0.81, 95%CI 0.41-1.58) and gastroschisis (aOR=2.93, 95%CI 0.97-8.88) were no longer significant in both crude and adjusted analyses. Maternal consumption of diclofenac was independently associated with a significant decrease in stillbirth (aOR=0.59, 95%CI 0.41-0.87). It is also interesting to note that diclofenac was the only sub-group analysis agreeing with the main analysis (exposure to at least one analgesic) on the significance of exposure association with increased incidence of neural tube defects (aOR=3.62, 95%CI 1.95-6.74) and hypospadias (aOR=1.49, 95%CI 1.09-2.03) compared to unexposed pregnancies in adjusted models.

## Discussion

### Main Findings

Consumption of paracetamol, ibuprofen, aspirin and naproxen during pregnancy, either in combination or separately, was significantly associated with increased premature delivery, stillbirth, neonatal death, LBW, abnormal standardised birthweight score and more frequent admission to neonatal unit. Consumption of paracetamol alone was further associated with higher odds for APGAR score <7 at one and five minutes both in crude and adjusted analyses. There was a dramatic increase in the frequency of over-the-counter (non-prescription) analgesic use in pregnancies between 1985 and 2015, reaching 70.5% of women in the final decade of our study. This means that our findings are applicable far beyond the percentage (between 14% and 38%) ^25^ of pregnant women with underlying health deficits related to the adverse outcomes we report here.

Diclofenac use increased steeply from 2005 (Figure 2A), which reflects the change in Scottish legislation, leading to diclofenac becoming available without prescription in that year. Diclofenac use was associated with fewer adverse outcomes but showed increased risk of neural tube defects and hypospadias in male neonates. Furthermore, and surprisingly, exposure to diclofenac only was associated with significant decrease in the incidence of stillbirth. The reasons for such differences between the changes in neonatal outcomes following diclofenac consumption compared with those following use of the other NSAIDS are not clear. The proportion of women using diclofenac, especially in the last 7 years of our study makes it highly unlikely to be due to an underlying maternal condition and/or other compounds used in combination (e.g. prescriptions) by women taking diclofenac. It is possible that the drug could act directly on fetal development then this difference could also be due to structural and/or mechanistic differences of the compound compared to the other drugs. However, not enough is known about the specific mechanisms of action of the different analgesics studied to conclude further. Overall, comparing our main analysis with all three sub-analyses, it is evident that the most significant differences were observed when paracetamol was taken with at least one other analgesic. This is mostly due to the high number of pregnancies where paracetamol was used, comprising almost 55% of the exposed cases in the main analysis.

### Strengths and Limitations

A major strength of the present study is the large cohort of 151,141 pregnancies over a 30-year study period from 1985 until 2015, using a robust data source AMND. This is one of the largest cohorts used in studies examining the effects of analgesic use during pregnancy. The dataset contains high quality and consistent data from the geographically defined area of Aberdeen and surrounding district, in the North East of Scotland, UK. In addition, as Aberdeen Maternity Hospital is the only maternity hospital serving the area, over 95% of pregnancies in the area are included in the dataset, considerably minimizing the risk for selection bias. We were able to analyse maternal consumption data of the five most commonly used analgesics available over-the-counter in the UK and most countries, which is not matched in the current literature. The nature of our data allowed for the analysis of analgesics consumed alone or in combination, unlike most existing studies, and this gives our study the added strength of better reflecting real-life consumption patterns ^26,27^. We were able to adjust for important confounding factors, relevant to each analysed outcome. Adjustment for maternal deprivation also allowed us to further account for potential unmeasured factors that can influence maternal and neonatal health, which is a major strength of our analysis compared to most studies.

A potential concern was that women were probably using analgesics to treat some inherent medical condition which in turn could have been the mediating factor for adverse outcomes. However, since these medications are widely available without prescription, this is unlikely to be a factor that affects the findings of this study. This is especially the case during the “diclofenac analysis” covering 2005-2015, where this study presents results on multiple neonatal outcomes for the given cohort. In this way we offer a comprehensive approach to the exploration of associations with *in utero* analgesic exposure rather than only focusing on a single outcome of interest. Our data were based on medical notes; however, over-the-counter consumption is self-reported, and details on the timing, dosage, product type (single-ingredient vs combination) and administration type were not available in the database. Complete case analyses were performed ignoring pregnancies with missing data in the covariates, however due to the low number of missing data there is little chance that this might have affected the validity of our results. Compared to our cohort size, there were, overall, very few cases of cryptorchidism, neural tube defects, amniotic band defects, hypospadias and gastroschisis, resulting in potentially underpowered statistical analyses to detect a difference for these outcomes. Our study only considered neonatal health outcomes and follow-up of the offspring was not available at this time.

### Interpretation

Previous literature has considered fewer outcomes with fewer analgesic combinations compared to our study. Consistent with our results, increased risk of preterm birth and miscarriage has been associated with analgesic consumption during pregnancy ^28–31^, while others reported no associations with miscarriage, stillbirth or preterm delivery ^20,28,29,32^. Similarly, increased risk for offspring cryptorchidism, hypospadias, neural tube defects, amniotic band defects and gastroschisis have been shown by many studies ^7–9,33–40^, although, again, a lack of associations with major birth defects have been reported ^13–17,41,42^. Compared to our analysis, all these studies used a smaller cohort, considered a shorter study time and there was frequent disagreement with respect to the choices of adjusted confounding factors. Another difference is that maternal questionnaires/interviews were frequently the method of choice to evaluate maternal consumption. Some of the studies reported increased risks for specific pregnancy trimesters which is something our study could not evaluate. Differences in study design and adjustment for different confounders might also account for the disagreement of our results that provide a more accurate assessment. Our study is one of the largest in terms of cohort size, duration, number of analgesics and range of outcomes included which might also contribute to differences compared to other studies.

The literature currently reports conflicting evidence, limiting our ability for definite decision-making. Over-the-counter analgesics are recommended to women by healthcare professionals in order to deal with pregnancy symptoms and other conditions. Policy-makers have taken a stand on the topic, either being reassuring about over-the-counter use during pregnancy or recommending caution when consumption is necessary ^43–46^. Different compounds can affect the mother and the fetus in a different way, and their combined use might worsen the risk for offspring ill health. This study demonstrates the need for additional research, before the field can be confidently directed towards one direction or the other.

Whether the associations we report result from flu, fever, rheumatological or inflammatory conditions, and/or combination with other prescribed medications or solely related to over-the-counter analgesics consumption is a matter of further research. Underlying health conditions could well influence the outcomes we see in this study, however, as these could be very different conditions it is biologically unlikely that they are responsible for the effects we observe here. Our study demonstrates an association of maternal over-the-counter analgesic consumption during pregnancy with adverse neonatal offspring outcomes. Future collaborative approaches such as an individual patient data meta-analysis that includes follow-up data on long-term outcomes during childhood and adulthood would significantly inform decision making. Going forward, uncovering the mechanisms of action and off target effects will also provide a solid foundation for the development of pregnancy-safe compounds. Finally, the findings present here suggest that diclofenac is associated with fewer changes in risk for the more frequent adverse outcomes although it is associated more with rarer, but severe, negative outcomes, including neural tube defects. Diclofenac may have a lower risk for the main adverse neonatal outcomes reported for paracetamol. However, it should be noted that our study is not designed to specifically test differences in level of risk between the analgesics included. Therefore, it should be emphasised that this does not mean that the authors are stating that diclofenac is preferable to paracetamol.

## Conclusions

Pain control is currently a therapeutic priority during pregnancy. Our findings of increased risk of adverse health outcomes for the offspring following at least first trimester maternal use of readily available over-the-counter analgesics are crucial to information for the management of pain during pregnancy.

## Data Availability

All data used in this study come from the AMND database. Approval by the Databank Steering Committee is required for access to the data, which has been granted for the purpose of this analysis.

## Acknowledgements

N/A

## Contribution to Authorship

AZ, SB and PAF contributed to the conception, design and coordination of the research. EAR provided critical input in the design and planning of statistical analysis. AZ conducted the statistical analysis and prepared the manuscript, figures and tables. AZ, SB, PAF, RTM and DCH substantially contributed to the analysis and interpretation of the work. All authors contributed to critical discussion of intellectual content, development and review/approval of the final manuscript version.

## Ethics Statement

The AMND dataset used in this study was fully anonymised, therefore there was no requirement for ethical approval. The North of Scotland Research Ethics Service has devolved Caldicott approval to the Chair of the AMND steering committee. Approval to access and analyse data was obtained from the AMND steering Committee (AMND 004/2018).

---

**Figure S1.**
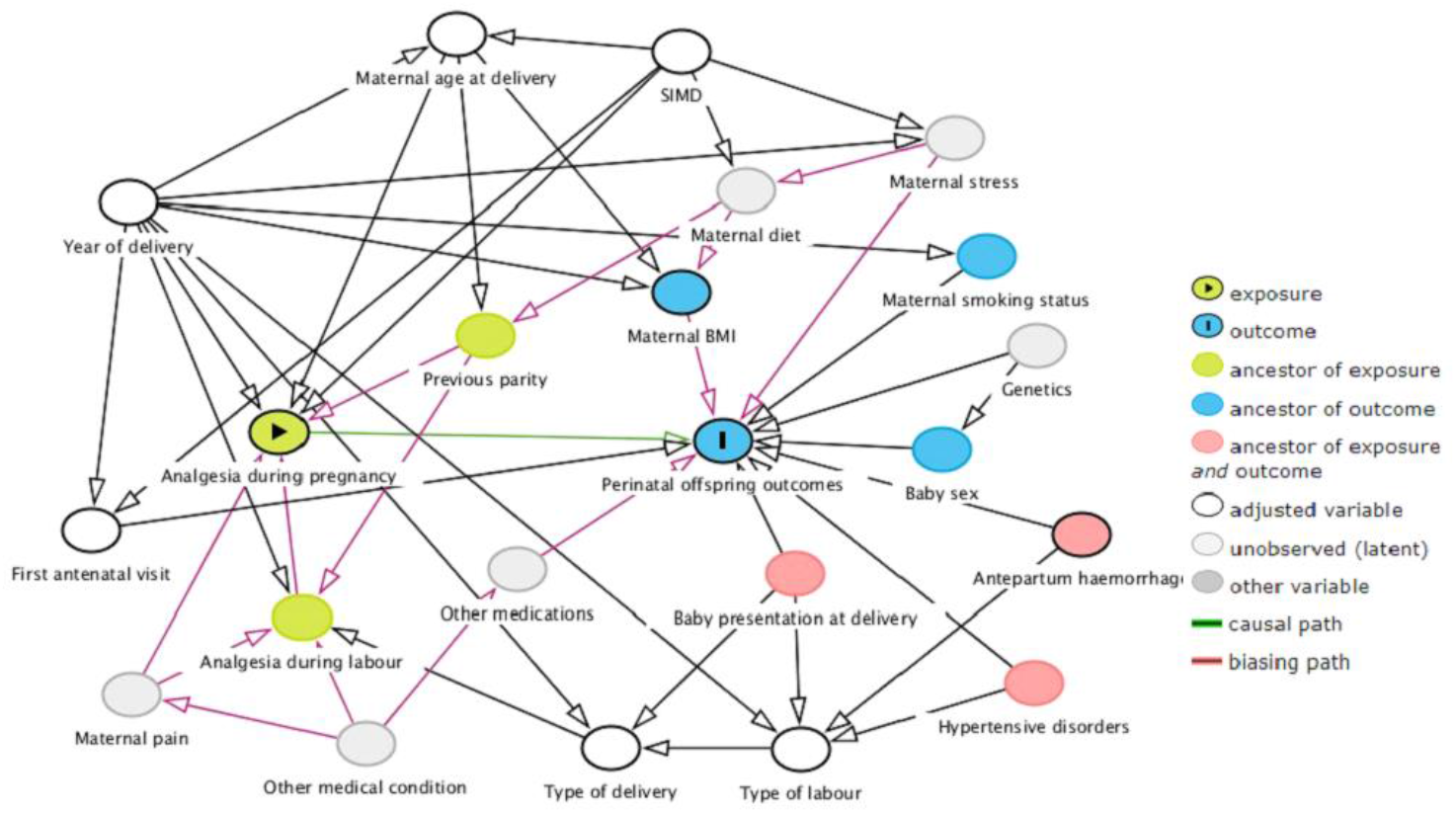
Directed acyclic graph (DAG) of exposure to outcome path and relevant measured and unmeasured biasing factors in our analysis.

